# Temporal Dynamics and Spectrum of Stroke Across Cancer Trajectories: A Multicenter Study of 190,000 Cancer Survivors

**DOI:** 10.64898/2026.10.12.26365360

**Authors:** Yasufumi Gon, Toshitaka Morishima, Tomohiro Kawano, Tsutomu Sasaki, Isao Miyashiro

## Abstract

**Background:** Stroke risk is increased in cancer patients, but variations by stroke type, cancer characteristics, and treatment timing remain incompletely characterized. We investigated stroke incidence after cancer diagnosis and temporal patterns of ischemic stroke around treatments.

**Methods:** We conducted a retrospective multicenter cohort study using linked cancer registry and administrative data from 68 cancer hospitals in Osaka, Japan. Patients diagnosed between 2019 and 2022 were followed for up to 1 year. In diagnosis-based analyses, standardized incidence ratios (SIRs) compared stroke incidence with the general population. In treatment-based analyses, ischemic stroke incidence rates and adjusted incidence rate ratios (IRRs) were estimated from 14 days before through 365 days after cancer treatments (surgery, chemotherapy, or radiotherapy).

**Results:** Among 190,471 patients, 1660 strokes occurred: 1296 (78.0%) ischemic strokes, 315 (19.0%) intracerebral hemorrhages, and 49 (3.0%) subarachnoid hemorrhages. In diagnosis-based analyses, the overall stroke SIR was 2.13 (95% CI, 2.03–2.24), with a 1.01% 1-year cumulative incidence. SIRs were 2.37 (95% CI, 2.24–2.50), 1.72 (95% CI, 1.53–1.92), and 1.22 (95% CI, 0.91–1.62) for ischemic stroke, intracerebral hemorrhage, and subarachnoid hemorrhage, respectively. SIRs peaked immediately after diagnosis and declined thereafter. Excess risk persisted through 180 days for ischemic stroke, 30 days for intracerebral hemorrhage, and 7 days for subarachnoid hemorrhage. In treatment-based analyses, ischemic stroke incidence increased transiently around treatments. After surgery, the adjusted IRR peaked during days 0–7 (12.27 [95% CI, 8.50–17.71]) and remained elevated through days 31– 90 versus days 181–365. A postoperative increase was observed even in early-stage disease. For chemotherapy, incidence was relatively high before treatment initiation.

**Conclusions:** Stroke risk after cancer diagnosis varied by stroke type, cancer characteristics, and time since diagnosis. Ischemic stroke incidence increased transiently around cancer treatment, including an early postoperative increase in patients with early-stage disease. These findings identify periods and patient groups that may warrant heightened clinical vigilance.

## INTRODUCTION

Advances in cancer care and population aging have led to an increasing number of cancer survivors.^1^ Cancer and stroke share risk factors such as smoking and diabetes mellitus.^2,3^ Cancer-associated coagulation abnormalities and cancer treatments, including surgery and chemotherapy, may further alter the risk of both thrombotic and hemorrhagic events.^4,5^ Consequently, patients with cancer are at increased risk of stroke. Given that the magnitude and temporal pattern of this risk vary by cancer type, clinical stage, disease activity, and treatment timing, characterizing these variations is essential for improving survivorship care.

The reported risk of stroke among patients with cancer varies across studies. A Swedish cohort study reported that patients with cancer had approximately 1.2-fold and 1.4-fold higher risks of ischemic and hemorrhagic stroke, respectively, than the general population.^6^ Stroke risk varies by cancer type, with particularly high risks reported among patients with brain or other central nervous system (CNS) tumors, pancreatic cancer, and uterine cancer.^7^ Lung cancer and hematologic malignancies were also associated with an increased risk of ischemic stroke (IS), which was particularly pronounced immediately after cancer diagnosis.^8,9^ A meta-analysis estimated the 1-year cumulative incidences of IS and intracerebral hemorrhage (ICH) after cancer diagnosis to be 1.3% and 0.3%, respectively.^10^ Despite these reports, few studies have comprehensively assessed the distribution and risks of IS, ICH, and subarachnoid hemorrhage (SAH) across diverse cancer types within the same cohort. Clarifying how the risk of each stroke type varies by cancer type and clinical stage, is important for characterizing the heterogeneous risk of stroke among patients with cancer.

Although cancer-related IS is often associated with advanced disease, it has also been reported in patients with early-stage cancer.^11^ Surgery may transiently increase stroke risk through physiological stress and hemostatic alterations,^12^ and pulmonary vein stump thrombosis after lung cancer resection may also cause systemic thromboembolism.^13^ Nevertheless, the temporal pattern of IS around cancer treatment remains poorly characterized, particularly whether its perioperative incidence varies by clinical stage.

This study aimed to characterize the risk of stroke after cancer diagnosis. We evaluated overall stroke and individual stroke types, including IS, ICH, and SAH, according to cancer type and clinical stage and examined temporal changes in IS incidence before and after cancer treatment.

## METHODS

### ETHICS APPROVAL AND CONSENT TO PARTICIPATE

This study complied with the Declaration of Helsinki and was approved by the Institutional Review Board of Osaka University Hospital (approval number: 22485). The need for written informed consent was waived because of the retrospective study design and an opt-out opportunity on the institutional website.

### DATA SOURCES

We conducted a retrospective multicenter cohort study to investigate the incidence of stroke among patients with cancer using data from the Cancer Registry-based Study on Cancer Care in Osaka (CanReCO) project. The CanReCO database was developed in collaboration with the Council for Coordination of Designated Cancer Care Hospitals in Osaka and comprises hospital-based cancer registry data linked to administrative data collected under Japan’s Diagnosis Procedure Combination (DPC) system. The DPC system is a standardized patient classification framework introduced by the Japanese government in 2003 to promote the standardization and evaluation of medical care and the appropriate allocation of healthcare resources.^14^ Unlike standalone cancer registry data used in our previous studies^15-18^, the linked CanReCO database includes information on noncancer diagnoses, medications, procedures, and other clinical factors, enabling analyses of incident noncancer conditions, including stroke, in relation to detailed patient and cancer characteristics.

Data were obtained from 68 hospitals in Osaka Prefecture that were currently or formerly designated cancer care hospitals.^19-21^ Cancer registry records for patients diagnosed between January 2019 and December 2022 were linked at the patient level to DPC records available from November 2018 through June 2023. Comorbidities present at admission and complications occurring after admission are separately recorded in the DPC data using International Classification of Diseases, Tenth Revision (ICD-10) codes, and the validity of these records has been reported previously.^22^

### STUDY POPULATION

The study population comprised patients diagnosed with cancer between January 2019 and December 2022 who were diagnosed and treated at a participating hospital, including those referred after diagnosis elsewhere. For patients with multiple primary cancers, only the first primary cancer was included as the index cancer, and records corresponding to subsequent primary cancers were excluded. We also excluded patients who had no follow-up information after cancer diagnosis, had no baseline DPC information available within 60 days before or after diagnosis, or had duplicate records. The patient selection process is shown in **Supplementary Figure S1**.

### CLINICAL VARIABLES

The CanReCO database provided variables derived from both the cancer registry and DPC records. Cancer registry data obtained at the time of cancer diagnosis included sex, age, cancer type, clinical stage, and adenocarcinoma. The registry also provided information on initial cancer treatment, including surgery, chemotherapy, and radiotherapy. DPC-derived baseline variables included body mass index (BMI), smoking history, comorbidities (hypertension, diabetes mellitus, dyslipidemia, atrial fibrillation, history of stroke, heart failure, and chronic kidney disease), and use of antiplatelet and anticoagulant agents. Comorbidities and other baseline clinical variables were ascertained from DPC records within 60 days before or after cancer diagnosis. Medication use was defined according to prescriptions recorded before cancer diagnosis.

Clinical stage was classified according to the Union for International Cancer Control TNM Classification of Malignant Tumors as stage 0, I, II, III, IV, unstaged, or not applicable.^23,24^ The ‘not applicable’ category included hematologic malignancies, such as leukemia and multiple myeloma; primary CNS tumors; and cancers of unknown primary site. Cancer treatment was defined as the first course of treatment administered for the index cancer according to the initial treatment plan. For hematologic malignancies, initial treatment included all therapies administered until the achievement of initial remission. Cancer surgery included open and minimally invasive procedures performed using optical instruments, such as thoracoscopic and laparoscopic surgery, but excluded endoscopic treatment alone. The International Classification of Diseases for Oncology, Third Edition, was used to code the cancer (**Supplemental Table S1**).

### OUTCOMES AND FOLLOW-UP

The observation period was from January 2019 through June 2023. The primary outcome was the first occurrence of stroke after cancer diagnosis. Patients were followed from the date of cancer diagnosis (the index date) until stroke occurrence, diagnosis of a second primary cancer, loss to follow-up, death, one year after cancer diagnosis, or June 30, 2023, whichever came first. Death was defined as death from any cause.

Stroke events, including those recorded as complications occurring after admission, were identified using ICD-10 codes together with DPC information on stroke-related procedures and care, including intravenous thrombolysis, mechanical thrombectomy, and stroke care unit management. The composite stroke outcome comprised IS, ICH, and SAH. The corresponding ICD-10 codes were I63 and I69.3 for IS, I61 and I69.1 for ICH, and I60 and I69.0 for SAH.

### STATISTICAL ANALYSIS

Baseline characteristics were summarized as medians with interquartile ranges (IQRs; Q1–Q3) for continuous variables and count with percentage for categorical variables.

To compare stroke risk among patients with cancer with that in the general population, standardized incidence ratios (SIRs) and their 95% confidence intervals (CIs) were calculated as the ratio of the observed to the expected number of stroke events. The observed number of events was obtained from the CanReCO database. The expected number was calculated by summing the products of the person-time contributed by the study cohort in each sex- and age-specific stratum and the corresponding national estimate stroke incidence, derived from population-based data collected in Shiga Prefecture, Japan.^25^ In accordance with the age categories available for the reference incidence rates, patients were stratified into the following age groups for the SIR calculations: <35, 35–44, 45–54, 55–64, 65–74, 75–84, and ≥85 years. SIRs were calculated for overall stroke and separately for IS, ICH, and SAH. SIRs were estimated for the entire follow-up period and separately for days 0–7, 8–14, 15–30, 31–90, 91– 180, and 181–365 after cancer diagnosis to characterize the temporal pattern of stroke risk. Analyses were also stratified by cancer type and clinical stage at diagnosis. The 1-year cumulative incidence of stroke was estimated using a competing-risk approach, treating death as a competing event.

To investigate the incidence of IS before and after cancer treatment, we identified patients who underwent cancer surgery, chemotherapy, or radiotherapy. For each treatment-specific analysis, the index date was defined as the date of surgery or the initiation date of chemotherapy or radiotherapy. Incidence rates of IS were calculated from 14 days before through 365 days after the index date. This period was divided into the following intervals: days −14 to −8, −7 to −1, 0–7, 8–14, 15–30, 31–90, 91–180, and 181–365. Adjusted incidence rate ratios (IRRs) and their 95% CIs for these intervals were estimated using Poisson regression models with log person-time as an offset, with days 181–365 serving as the reference interval. The models were adjusted for age, sex, BMI, hypertension, dyslipidemia, diabetes mellitus, history of stroke, atrial fibrillation, smoking status, heart failure, chronic kidney disease, use of antiplatelet agents, use of anticoagulants, clinical stage at cancer diagnosis, and adenocarcinoma histology. Robust standard errors clustered at the patient level were used to account for within-patient correlation. Among patients who underwent cancer surgery, incidence rates of IS were also calculated according to clinical stage at cancer diagnosis. Missing covariate data were handled using multiple imputation by chained equations, generating 10 imputed datasets. Estimates were pooled using Rubin’s rules.^26^

Statistical analyses were performed using R software (R Foundation for Statistical Computing, Vienna, Austria). All tests were two-sided, and a *P* value <0.05 was considered statistically significant.

### DATA Availability

The data that support the findings of this study are available from the corresponding author upon reasonable request.

## RESULTS

### STUDY POPULATION

Table 1 summarizes the baseline characteristics of the 190,471 patients included in the analysis. Overall, 46.3% were women, and patients aged 65-74 years constituted the largest age group. The median BMI was 22.4 kg/m² (Q1–Q3, 20.1–24.9). A history of smoking was reported in 48.4%, and hypertension was the most common comorbidity (30.8%). Antiplatelet and anticoagulant agents were prescribed to 5.8% and 3.8% of patients, respectively. Stage I was the most common stage at diagnosis (33.3%), followed by stage IV (18.5%). Cancer surgery, chemotherapy, and radiotherapy were performed in 52.8%, 34.8%, and 11.8% of patients, respectively. Colorectal cancer was the most common cancer type (16.6%), followed by lung (12.8%), gastric (10.9%), breast (9.0%), and prostate cancers (7.4%).

**Table 1.**
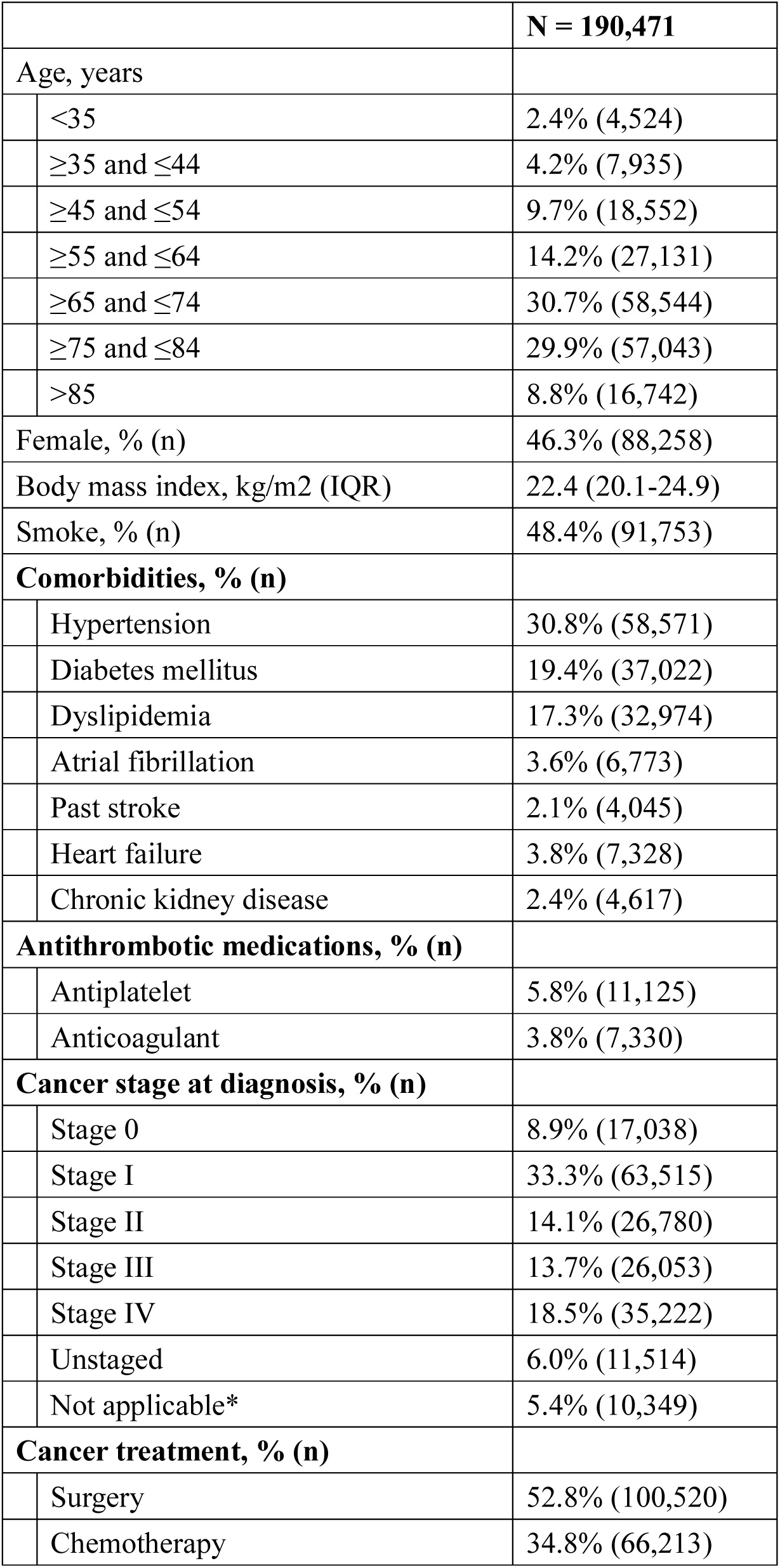
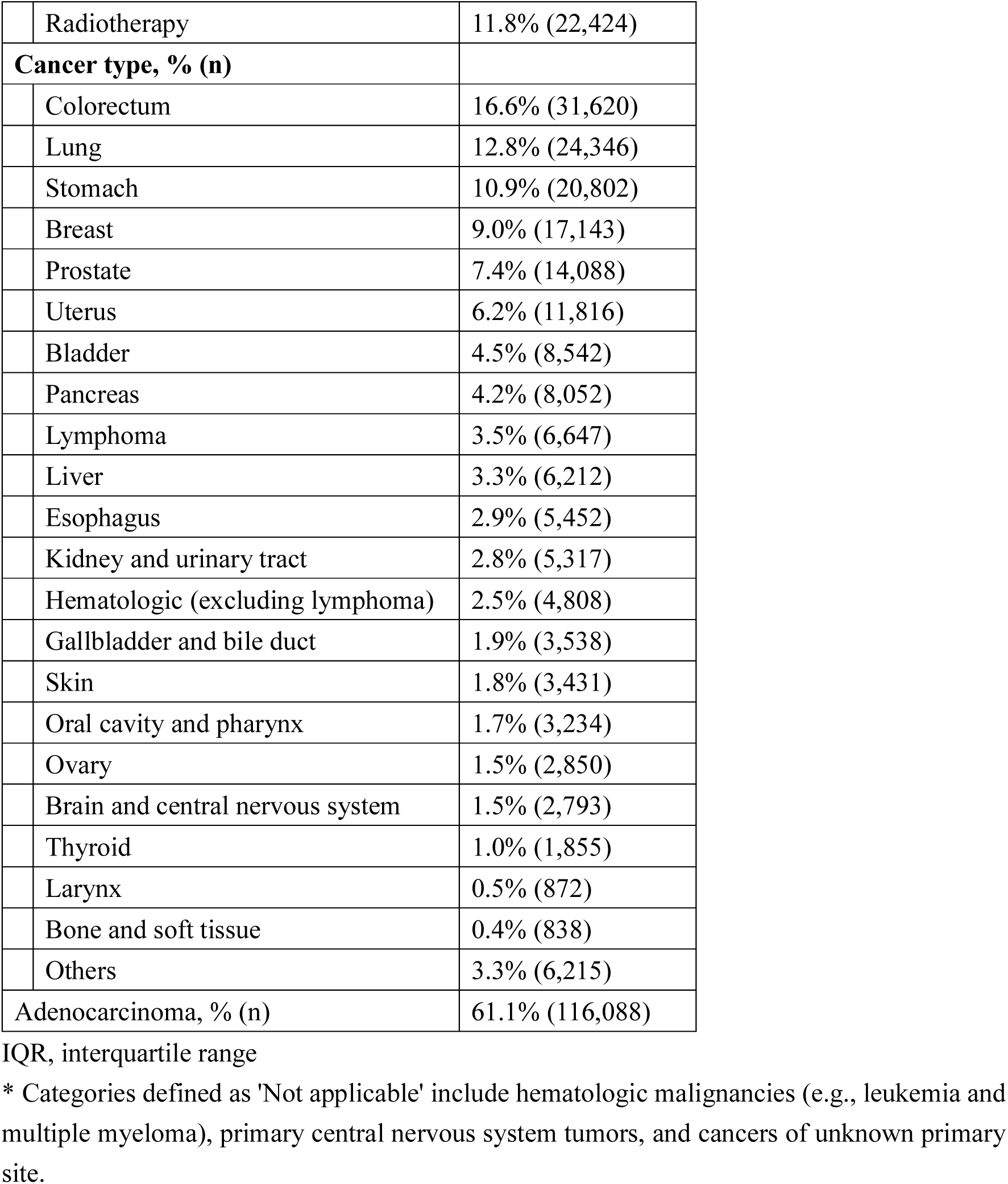
Baseline characteristics of the cohort.

### STROKE INCIDENCE AFTER CANCER DIAGNOSIS

**Table 2** summarizes stroke incidence after cancer diagnosis. The overall SIR for stroke was 2.13 (95% CI, 2.03–2.24), with an incidence rate of 12.34 per 1,000 person-years and a 1-year cumulative incidence of 1.01%. Of the stroke events, 78.0% were IS, 19.0% were ICH, and 3.0% were SAH. SIRs were highest immediately after cancer diagnosis and declined thereafter. The excess risk persisted through 180 days for IS and 30 days for ICH but was confined to days 0–7 for SAH. The corresponding cumulative incidences were 0.79% for IS, 0.20% for ICH, and 0.03% for SAH.

**Table 2.** Standardized incidence ratio and incidence rate of stroke after cancer diagnosis.

| Outcome |  | Period after cancer diagnosis, days | Observed, n | Expected, n | SIR (95% CI) | IR per 1000 PY (95% CI) |
| --- | --- | --- | --- | --- | --- | --- |
| All stroke | Overall |  | 1,660 | 779.1 | 2.13 (2.03-2.24) | 12.34 (11.75-12.95) |
|  |  | 0-7 | 471 | 22.5 | 20.95 (19.10-22.93) | 130.16 (118.67-142.47) |
|  |  | 8-14 | 152 | 21.9 | 6.93 (5.87-8.12) | 42.80 (36.26-50.17) |
|  |  | 15-30 | 162 | 48.0 | 3.38 (2.88-3.94) | 20.61 (17.56-24.04) |
|  |  | 31-90 | 297 | 162.1 | 1.83 (1.63-2.05) | 10.89 (9.69-12.20) |
|  |  | 91-180 | 278 | 214.6 | 1.30 (1.15-1.46) | 7.49 (6.64-8.43) |
|  |  | 181-365 | 300 | 309.9 | 0.97 (0.86-1.08) | 5.44 (4.84-6.10) |
| IS | Overall |  | 1296 | 546.6 | 2.37 (2.24-2.50) | 9.63 (9.12-10.17) |
|  |  | 0-7 | 353 | 15.9 | 22.22 (19.96-24.67) | 97.55 (87.64-108.28) |
|  |  | 8-14 | 123 | 15.5 | 7.94 (6.60-9.48) | 34.63 (28.78-41.32) |
|  |  | 15-30 | 132 | 33.8 | 3.90 (3.26-4.63) | 16.80 (14.05-19.92) |
|  |  | 31-90 | 248 | 114.0 | 2.17 (1.91-2.46) | 9.09 (8.00-10.30) |
|  |  | 91-180 | 216 | 150.5 | 1.43 (1.25-1.64) | 5.82 (5.07-6.65) |
|  |  | 181-365 | 224 | 216.8 | 1.03 (0.90-1.18) | 4.06 (3.55-4.63) |
| ICH | Overall |  | 315 | 183.6 | 1.72 (1.53-1.92) | 2.34 (2.09-2.62) |
|  |  | 0-7 | 101 | 5.2 | 19.28 (15.71-23.43) | 27.91 (22.73-33.92) |
|  |  | 8-14 | 27 | 5.1 | 5.28 (3.48-7.68) | 7.60 (5.01-11.06) |
|  |  | 15-30 | 25 | 11.2 | 2.23 (1.44-3.29) | 3.18 (2.06-4.70) |
|  |  | 31-90 | 41 | 38.1 | 1.08 (0.77-1.46) | 1.50 (1.08-2.04) |
|  |  | 91-180 | 57 | 50.6 | 1.13 (0.85-1.46) | 1.54 (1.16-1.99) |
|  |  | 181-365 | 64 | 73.4 | 0.87 (0.67-1.11) | 1.16 (0.89-1.48) |
| SAH | Overall |  | 49 | 40.1 | 1.22 (0.91-1.62) | 0.36 (0.27-0.48) |
|  |  | 0-7 | 17 | 1.1 | 15.46 (9.00-24.75) | 4.70 (2.74-7.52) |
|  |  | 8-14 | 2 | 1.1 | 1.86 (0.22-6.70) | 0.56 (0.07-2.03) |
|  |  | 15-30 | 5 | 2.4 | 2.11 (0.68-4.91) | 0.64 (0.21-1.48) |
|  |  | 31-90 | 8 | 8.2 | 0.98 (0.42-1.93) | 0.29 (0.13-0.58) |
|  |  | 91-180 | 5 | 11.0 | 0.45 (0.15-1.06) | 0.13 (0.04-0.31) |
|  |  | 181-365 | 12 | 16.3 | 0.74 (0.38-1.29) | 0.22 (0.11-0.38) |

SIR, standardized incidence ratio; CI, confidence interval; IR, incidence rate; PY, person-years;
IS, ischemic stroke; ICH, intracerebral hemorrhage; SAH, subarachnoid hemorrhage

**Figure 1** presents the SIRs for each stroke type by cancer type. Brain and CNS tumors had the highest SIRs for all 3 stroke types: SIRs for IS, ICH, and SAH were 14.18 (95% CI, 10.97–18.04), 44.69 (95% CI, 35.32–55.77), and 7.99 (95% CI, 2.18–20.46), respectively. High SIRs were also observed for IS in ovarian (9.09 [95% CI, 5.99–13.23]) and pancreatic (6.40 [95% CI, 5.32–7.62]) cancers. For both ICH and SAH, the second-highest SIRs were observed in other hematologic malignancies, after brain and CNS tumors. Detailed estimates are provided in **Supplementary Table S2.**

**Figure 1.**
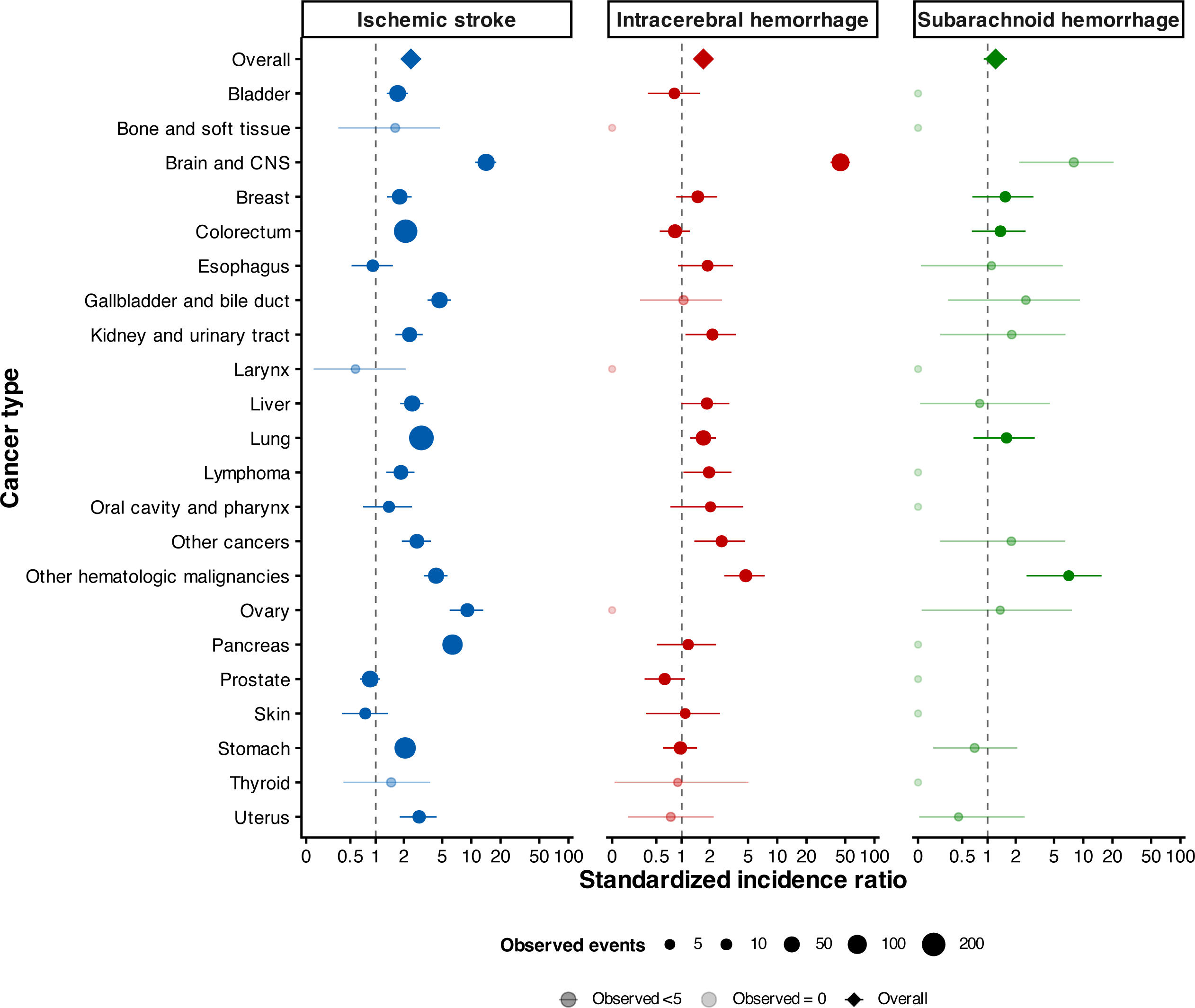
Standardized incidence ratios for stroke type according to cancer type. Standardized incidence ratios (SIRs) and their 95% confidence intervals (CIs) are shown for ischemic stroke, intracerebral hemorrhage, and subarachnoid hemorrhage according to cancer type. Circles represent cancer type–specific SIRs, and horizontal lines represent 95% CIs. The size of each circle is proportional to the number of observed events. Diamonds represent the overall estimates. Lighter symbols indicate estimates based on fewer than 5 or no observed events. The vertical dashed line indicates an SIR of 1.0. For ischemic stroke, the highest SIRs were observed for brain and central nervous system tumors (SIR, 14.18 [95% CI, 10.97–18.04]), ovarian cancer (SIR, 9.09 [95% CI, 5.99–13.23]), and pancreatic cancer (SIR, 6.40 [95% CI, 5.32–7.62]). For intracerebral hemorrhage, the highest SIRs were observed for brain and central nervous system tumors (SIR, 44.69 [95% CI, 35.32–55.77]) and other hematologic malignancies excluding lymphoma (SIR, 4.73 [95% CI, 2.85–7.39]). For subarachnoid hemorrhage, the corresponding estimates were highest for brain and central nervous system tumors (SIR, 7.99 [95% CI, 2.18–20.46]) and other hematologic malignancies excluding lymphoma (SIR, 7.09 [95% CI, 2.60–15.44]). CI, confidence interval; CNS, central nervous system; SIR, standardized incidence ratio.

**Figure 2** shows the corresponding SIRs according to clinical stage at cancer diagnosis. The SIR for IS generally increased with advancing cancer stage, and ICH risk was elevated in patients with stage III or IV disease, whereas no consistent stage-dependent pattern was observed for SAH. Detailed estimates are provided in **Supplementary Table S3**.

**Figure 2.**
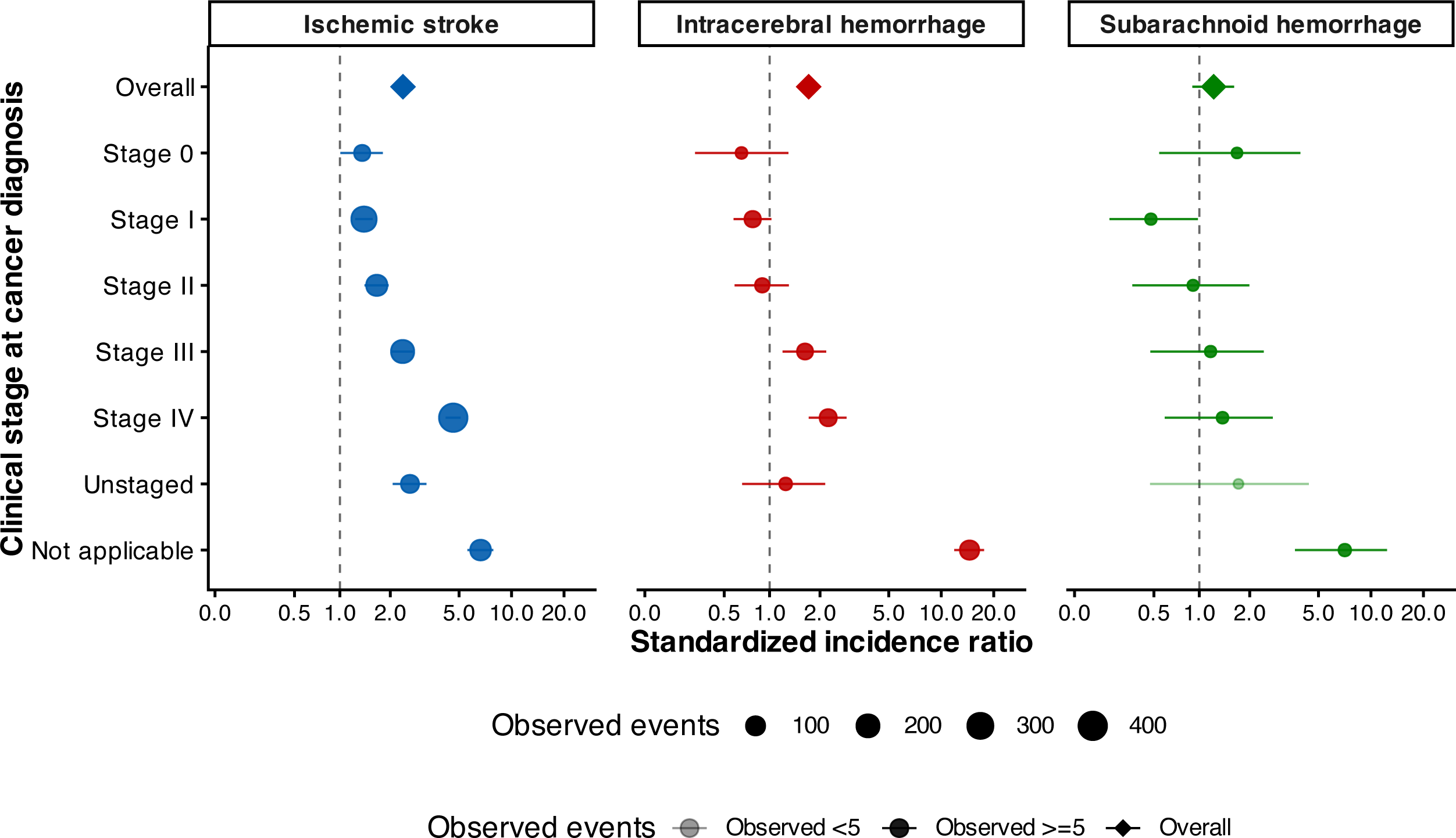
Standardized incidence ratios for stroke type according to clinical stage at cancer diagnosis. Standardized incidence ratios (SIRs) and their 95% confidence intervals (CIs) are shown for ischemic stroke, intracerebral hemorrhage, and subarachnoid hemorrhage according to clinical stage at cancer diagnosis. Circles represent stage-specific SIRs, and horizontal lines represent 95% CIs. The size of each circle is proportional to the number of observed events. Diamonds represent the overall estimates. Lighter symbols indicate estimates based on fewer than 5 observed events. The vertical dashed line indicates an SIR of 1.0. For ischemic stroke, the SIR generally increased with advancing clinical stage and was highest in patients with stage IV disease. For intracerebral hemorrhage, elevated SIRs were observed in patients with stage III or IV disease. Particularly high estimates for ischemic stroke and intracerebral hemorrhage were observed in patients for whom clinical stage was not applicable. No consistent stage-dependent pattern was observed for subarachnoid hemorrhage. CI, confidence interval; SIR, standardized incidence ratio.

### RISK OF IS BEFORE AND AFTER CANCER TREATMENT

**Figure 3** shows the incidence rates and adjusted IRRs for IS according to time relative to cancer treatment, with days 181–365 after treatment used as the reference period. Detailed estimates are provided in **Supplementary Table S4**. After cancer surgery, the incidence rate increased sharply, peaked during days 0–7, and progressively declined thereafter. The adjusted IRR was highest during days 0–7 (12.27; 95% CI, 8.50–17.71) and remained significantly elevated through days 31–90, whereas no significant elevation was observed during days 91–180.

**Figure 3.**
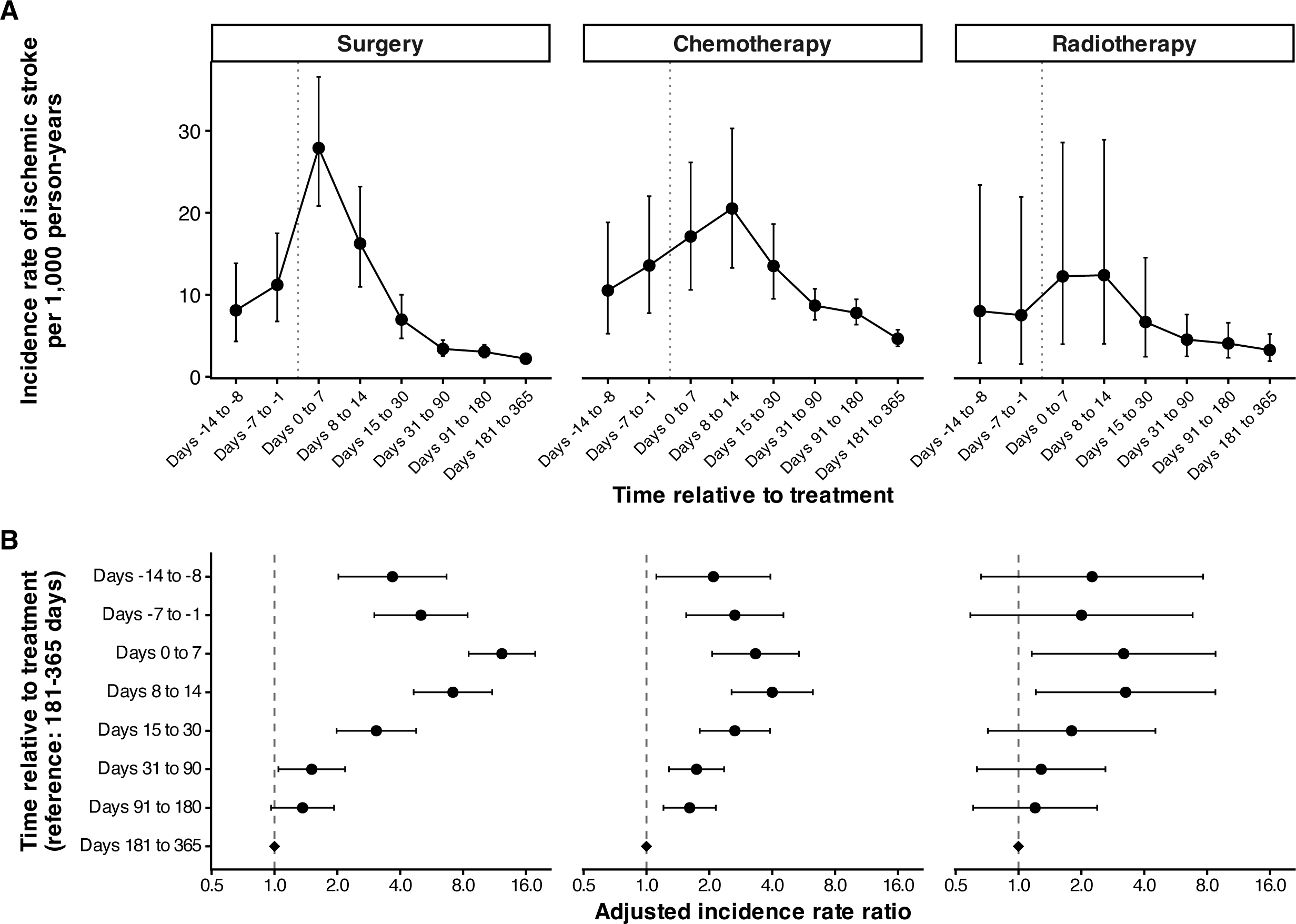
Incidence of ischemic stroke according to time relative to cancer treatment. Panel A shows the incidence rates of ischemic stroke per 1,000 person-years and their 95% confidence intervals during defined intervals before and after cancer surgery, chemotherapy, and radiotherapy. The vertical dotted line indicates the index treatment date. Panel B shows adjusted incidence rate ratios and their 95% confidence intervals for the corresponding intervals, with days 181–365 after treatment used as the reference period. Circles represent point estimates, horizontal lines represent 95% confidence intervals, and diamonds represent the reference estimates. The vertical dashed line in Panel B indicates an incidence rate ratio of 1.0. The models were adjusted for age, sex, body mass index, hypertension, dyslipidemia, diabetes mellitus, history of stroke, atrial fibrillation, smoking status, heart failure, chronic kidney disease, use of antiplatelet agents, use of anticoagulants, cancer stage at diagnosis, and adenocarcinoma histology. Estimates were pooled across 10 multiply imputed datasets using Rubin’s rules. CI, confidence interval; IRR, incidence rate ratio.

Among patients receiving chemotherapy, the incidence rate began increasing before treatment, peaked during days 8–14 after treatment, and subsequently declined. The adjusted IRRs were significantly elevated before treatment and remained elevated through days 91–180 after treatment, with the highest estimate observed during days 8–14 (adjusted IRR, 4.00; 95% CI, 2.56–6.26).

Among patients receiving radiotherapy, the incidence rate was highest during the first 14 days after treatment and decreased thereafter. The adjusted IRRs were significantly elevated during both days 0–7 (3.19; 95% CI, 1.16–8.78) and days 8–14 (3.26; 95% CI, 1.21–8.77). Estimates for the remaining intervals had wide confidence intervals and did not differ significantly from the reference period.

### INCIDENCE OF IS AROUND CANCER SURGERY ACCORDING TO CANCER **STAGE**

**Figure 4** shows the incidence rates of IS before and after cancer surgery according to clinical stage at cancer diagnosis. Among patients with stage I–IV disease, the incidence rate generally peaked during the first 7 days after surgery and subsequently declined. A postoperative peak appeared more prominent in patients with stage I disease than in those with stage II-IV disease. In patients with stage III or IV disease, the incidence rate was already elevated during the 14 days preceding surgery, with a further increase immediately after surgery. By days 181–365, the incidence rates were substantially lower across all stages. No clear temporal pattern was observed for stage 0 because of the small number of events, as reflected by the wide confidence intervals. Confidence intervals were also wide for several perioperative estimates, particularly in patients with advanced-stage disease.

**Figure 4.**
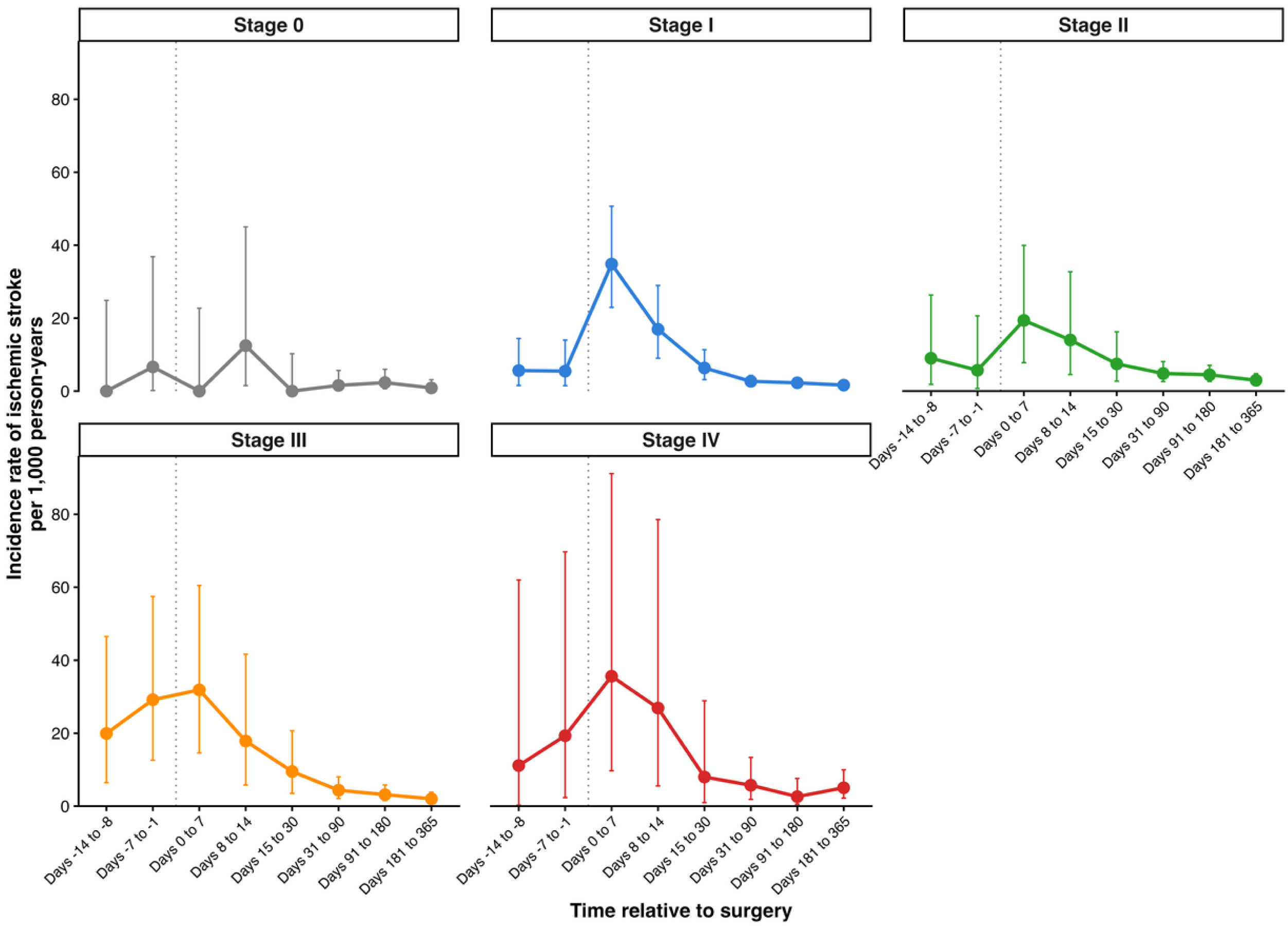
Incidence of ischemic stroke before and after cancer surgery according to clinical stage at cancer diagnosis. Incidence rates of ischemic stroke per 1,000 person-years and their 95% confidence intervals are shown for patients with stage 0–IV cancer during the following intervals relative to cancer surgery: days −14 to −8, days −7 to −1, days 0–7, days 8–14, days 15–30, days 31–90, days 91–180, and days 181–365. Points represent incidence-rate estimates, and vertical error bars represent 95% confidence intervals. The vertical dotted line indicates the index surgery date. A common y-axis scale was applied to all panels to facilitate comparisons across cancer stages; its upper limit was determined from the maximum upper 95% confidence limit among patients with stage IV disease. Upward-pointing symbols indicate confidence limits exceeding the displayed range. CI, confidence interval.

## DISCUSSION

In this large multicenter cohort of 190,471 patients with cancer, stroke risk was approximately twice that in the general population and was highest immediately after cancer diagnosis. The magnitude and duration of the excess risk varied by stroke type, cancer type, and clinical stage. A key contribution of this study was the characterization of IS incidence around cancer treatment. IS incidence increased transiently around cancer surgery, chemotherapy, and radiotherapy, with distinct temporal patterns across treatment modalities. The increase was most pronounced immediately after surgery and was observed not only in patients with stage III or IV disease but also in those with early-stage disease.

### CANCER TREATMENT AND IS INCIDENCE

A distinctive feature of this study was the evaluation of IS incidence before and after cancer treatment. Although transient increases were observed across treatment modalities, the increase was most pronounced immediately after cancer surgery. Several factors may have contributed to this finding. The elevated postoperative risk of IS after surgery for brain tumors and lung cancer—potentially related to pulmonary vein stump thrombosis in the latter—may have contributed to the overall estimate.^27,28^ In addition, surgical stress and the associated inflammatory and hemostatic responses may promote thrombosis, while the temporary interruption of antiplatelet or anticoagulant therapy may further increase perioperative risk.^12^ Although routine heparin bridging during interruption of oral anticoagulation has not reduced perioperative arterial thromboembolism and increases bleeding in general populations^29^, its potential benefit in selected patients undergoing cancer surgery who are at particularly high thrombotic risk remains uncertain. Although detailed information on the perioperative interruption, resumption, and bridging of antithrombotic therapy was unavailable, the marked postoperative peak observed in this study supports further investigation of individualized perioperative antithrombotic strategies. The subsequent decline in IS incidence may reflect the resolution of acute perioperative effects and, in patients successfully treated for cancer, a reduction in tumor burden accompanied by improvement in cancer-associated activation of coagulation.

### CANCER TYPE-SPECIFIC PATTERNS

Brain and CNS tumors had significantly elevated SIRs for all stroke types. This increased risk may primarily reflect the local effects of intracranial tumors and their treatment. Tumor-related compression or invasion of cerebral vessels and perioperative vascular injury may contribute to IS, particularly after neurosurgery.^27,30^ Disruption of cerebral tissue and vasculature, fragile tumor-associated vessels, intratumoral hemorrhage, and treatment-related complications may contribute to ICH.^27,30^ Similar local vascular and treatment-related factors may also increase the risk of SAH^27^, although the small number of SAH events limits mechanistic interpretation.

The elevated SIRs for IS in pancreatic and ovarian cancers may partly reflect the cancer-associated activation of coagulation with these malignancies.^31,32^ Experimental studies have shown that pancreatic cancer–derived tissue factor and tissue factor–positive extracellular vesicles promote coagulation and thrombosis, whereas ovarian clear cell carcinoma produces tissue factor–rich extracellular vesicles and induces systemic activation of coagulation.^33,34^ These prothrombotic abnormalities may have contributed to the elevated risk of IS observed in both malignancies. Lung cancer accounted for the largest number of IS events. In addition to cancer-associated activation of coagulation, thrombus formation at the pulmonary vein stump following lung resection may contribute to IS in some surgically treated patients.^28^ For ICH, a high SIR was observed among patients with other hematologic malignancies. Hematologic malignancies can cause complex hemostatic abnormalities, including activation of coagulation, hyperfibrinolysis, and thrombocytopenia.^35,36^ The SIR for SAH was also elevated in this group, possibly reflecting similar hemostatic abnormalities.^35,36^ However, the small number of events warrants cautious interpretation.

### STAGE-SPECIFIC PATTERNS

The SIRs for IS and ICH tended to increase with advancing cancer stage. For IS, this pattern may reflect the activation of coagulation associated with greater tumor burden and more active disease.^37^ The elevated risk of ICH in patients with stage III or IV cancer may be related to several factors, including intensive anticancer treatment, treatment-related thrombocytopenia or coagulation abnormalities, and hemorrhage from brain metastases.^30,38^ In contrast, no consistent stage-dependent pattern was observed for SAH.

Elevated SIRs for all stroke types were also observed in patients for whom clinical stage was not applicable. This finding may partly reflect the composition of this group, which included a substantial proportion of brain and CNS tumors, hematologic malignancies, and other cancers to which conventional solid-tumor staging was not applicable. Because the stage-specific analyses did not account for differences in cancer type, treatment, and other patient characteristics, these findings should not be interpreted as demonstrating an independent effect of cancer stage on stroke risk.

### TEMPORAL PATTERNS AFTER CANCER DIAGNOSIS

Consistent with previous studies, stroke risk was markedly elevated immediately after cancer diagnosis.^10,39^ This early increase is likely multifactorial. Active cancer, activation of coagulation, and the initiation of diagnostic and therapeutic procedures are associated with the increased risk during this period.^10,39^ In addition, increased healthcare contact and the frequent use of brain imaging around the time of cancer diagnosis may have increased stroke detection. The excess risk of IS remained significantly elevated through 180 days after cancer diagnosis but was no longer significantly higher than that in the general population thereafter. During periods of high cancer activity, cancer-associated activation of coagulation and other prothrombotic factors may be particularly pronounced, contributing to the excess risk of IS during the first 6 months.^39^ Among patients who responded to cancer treatment, a reduction in tumor burden may have ameliorated activation of coagulation and reduced the risk of IS; conversely, patients with disease too advanced to receive or benefit from treatment may have died or otherwise left the risk set, resulting in the selective survival of patients at lower risk.

In contrast, the excess risks of ICH and SAH were confined to earlier periods. The concentration of these events shortly after diagnosis may partly reflect the high risks associated with brain and CNS tumors and hematologic malignancies, including intratumoral hemorrhage, thrombocytopenia, coagulation abnormalities, and complications of initial cancer treatment.^27,30^ However, the smaller numbers of hemorrhagic events reduced statistical precision; therefore, the absence of a significantly elevated SIR during later periods does not necessarily indicate that the risk had completely resolved.

### DISTRIBUTION OF STROKE TYPES AMONG PATIENTS WITH CANCER

We previously reported that IS, ICH, and SAH accounted for 76.1%, 20.2%, and 3.7% of stroke events among patients with cancer, respectively; however, that study was limited by its single-center design.^40^ In the present multicenter cohort, the corresponding proportions were 78.0%, 19.0%, and 3.0%, respectively, broadly consistent with our previous findings. This distribution was also similar to that reported in the Japan Stroke Data Bank and to the distribution observed in high-income countries.^41,42^ Although patients with cancer might be expected to have a disproportionately high frequency of IS because activation of coagulation promotes thrombosis, the proportion of IS in our cohort was broadly comparable to that in the general population. This represents an important finding and highlights the clinical relevance of both ischemic and hemorrhagic stroke among patients with cancer.

### LIMITATIONS

A major strength of this study was its large, multicenter design, comprising approximately 190,000 patients with cancer. Nevertheless, several limitations should be acknowledged. First, the reference estimates of stroke incidence were derived from population-based data collected in Shiga Prefecture. Geographic differences in stroke incidence between Shiga and Osaka may therefore have affected the expected event counts and SIR estimates. Second, stroke outcomes were identified from ICD-10 codes and DPC information without independent clinical adjudication. For stroke events recorded as complications occurring after admission, ascertainment incorporated information on stroke-related procedures or care. Although this approach likely improved specificity, it may have missed events managed without qualifying procedures or care, potentially resulting in underestimation of stroke incidence. Third, stroke events treated outside the participating hospitals were not captured, potentially leading to underestimation of stroke incidence. Such underascertainment may have been greater later in follow-up and may have partly contributed to the observed temporal decline. Finally, the study was conducted exclusively in Japan. Differences in population characteristics, cancer care, stroke epidemiology, and healthcare systems may limit the generalizability of our findings, and further studies in other countries and regions are warranted.

## CONCLUSION

Patients with cancer had an increased risk of stroke, particularly immediately after cancer diagnosis, with substantial variation by stroke type, cancer type, clinical stage, and treatment timing. IS incidence increased transiently around cancer treatment, including after surgery for both early- and advanced-stage cancer. These findings may help identify clinically relevant periods and patient groups for heightened stroke surveillance.

## ACKNOWLEDGMENTS

None

## AUTHORSHIP CONTRIBUTIONS

Contribution: Y.G., T.M., T.K., and I.M. designed the research and analyzed the data; Y.G. wrote the manuscript; T.M., T.K., T.S. and I.M. provided critical guidance on statistical procedures, and helped in writing the manuscript; and all authors read and approved the final manuscript.

## SOURCE OF FUNDING

This work was supported by Grants-in-Aid for Scientific Research of the Japan Society for the Promotion of Science (grant number: JP23K09713).

## POTENTIAL CONFLICTS OF INTEREST

Nothing to report.

## Abbreviations

BMI: body mass index
CanReCO: Cancer Registry-based Study on Cancer Care in Osaka
CI: confidence interval
CNS: central nervous system
DPC: Diagnosis Procedure Combination
ICD-10: International Classification of Diseases, Tenth Revision
ICH: intracerebral hemorrhage
IQR: interquartile range
IRR: incidence rate ratio
IS: ischemic stroke
SAH: subarachnoid hemorrhage
SIR: standardized incidence ratio

## Notes

### Competing Interest Statement

The authors have declared no competing interest.

### Author Declarations

Institutional Review Board of Osaka University Hospital

## REFERENCES

1. Wagle NS, Nogueira L, Devasia TP, et al. Cancer treatment and survivorship statistics, 2025. CA Cancer J Clin. 2025;75:308–340. doi: 10.3322/caac.70011.

2. Alhuneafat L, Guha A, Blaes A, Konety SH. Cancer and Cardiovascular Disease: Shared Risk Factors, Mechanisms, and Clinical Implications: JACC: CardioOncology State-of-the-Art Review. JACC CardioOncol. 2025;7:453–469. doi: 10.1016/j.jaccao.2025.07.001.

3. Koene RJ, Prizment AE, Blaes A, Konety SH. Shared Risk Factors in Cardiovascular Disease and Cancer. Circulation. 2016;133:1104–14. doi: 10.1161/CIRCULATIONAHA.115.020406.

4. Dardiotis E, Aloizou AM, Markoula S, et al. Cancer-associated stroke: Pathophysiology, detection and management (Review). Int J Oncol. 2019;54:779–796. doi: 10.3892/ijo.2019.4669.

5. Kawano T, Mackman N. Cancer patients and ischemic stroke. Thromb Res. 2024;237:155–162. doi: 10.1016/j.thromres.2024.03.019.

6. Zöller B, Ji J, Sundquist J, Sundquist K. Risk of haemorrhagic and ischaemic stroke in patients with cancer: a nationwide follow-up study from Sweden. Eur J Cancer. 2012;48:1875–83. doi: 10.1016/j.ejca.2012.01.005.

7. Strongman H, Gadd S, Matthews A, et al. Medium and long-term risks of specific cardiovascular diseases in survivors of 20 adult cancers: a population-based cohort study using multiple linked UK electronic health records databases. Lancet. 2019;394:1041–1054. doi: 10.1016/S0140-6736(19)31674-5.

8. Navi BB, Reiner AS, Kamel H, et al. Risk of Arterial Thromboembolism in Patients With Cancer. J Am Coll Cardiol. 2017;70:926–938. doi: 10.1016/j.jacc.2017.06.047.

9. Gon Y, Morishima T, Kawano T, et al. Arterial Thromboembolism in Japanese Patients With Cancer: Incidence, Predictors, and Survival Impact. JACC CardioOncol. 2024;6:283–297. doi: 10.1016/j.jaccao.2024.01.006.

10. Lun R, Roy DC, Hao Y, et al. Incidence of stroke in the first year after diagnosis of cancer-A systematic review and meta-analysis. Front Neurol. 2022;13:966190. doi: 10.3389/fneur.2022.966190.

11. Navi BB, Reiner AS, Kamel H, et al. Arterial thromboembolic events preceding the diagnosis of cancer in older persons. Blood. 2019;133:781–789. doi: 10.1182/blood-2018-06-860874.

12. Rautiola J, Björklund J, Zelic R, et al. Risk of Postoperative Ischemic Stroke and Myocardial Infarction in Patients Operated for Cancer. Ann Surg Oncol. 2024;31:1739–1748. doi: 10.1245/s10434-023-14688-6.

13. Ezeja L, Jaradat S, Qian J. Cancer treatment-related risk factors for stroke among patients with lung cancer: A systematic review and meta-analysis. Cancer Treat Res Commun. 2026;47:101140. doi: 10.1016/j.ctarc.2026.101140.

14. Hayashida K, Murakami G, Matsuda S, Fushimi K. History and profile of Diagnosis Procedure Combination (DPC): Development of a real data collection system for acute inpatient care in Japan. J Epidemiol. 2021;31:1–11. doi:10.2188/jea.JE20200288.

15. Gon Y, Zha L, Sasaki T, et al. Stroke mortality in cancer survivors: A population-based study in Japan. Thromb Res. 2023;222:140–148.

16. Gon Y, Zha L, Sasaki T, et al. Heart Disease Mortality in Cancer Survivors: A Population-Based Study in Japan. J Am Heart Assoc. 2023;12:e029967. doi: 10.1161/JAHA.123.029967.

17. Gon Y, Zha L, Morishima T, et al. Non-cancer-related Deaths in Cancer Survivors: A Nationwide Population-based Study in Japan. J Epidemiol. 2025;35:147–153. doi: 10.2188/jea.JE20240230.

18. Gon Y, Zha L, Kawano T, et al. Specific Cardiovascular Mortality in Cancer Survivors: A Nationwide Population-Based Cohort Study in Japan. J Am Heart Assoc. 2025;14:e037965. doi: 10.1161/JAHA.124.037965.

19. Miyazaki A, Kunimasa K, Morishima T, et al. Comparative effectiveness of immunotherapeutic regimens for untreated advanced non-small-cell lung cancer in real-world clinical practice: a Japanese cancer registry cohort study. ESMO Real World Data Digit Oncol. 2025;8:100137. doi: 10.1016/j.esmorw.2025.100137.

20. Kajiwara-Saito M, Morishima T, Ma C, Koyama S, Miyashiro I. Diagnosis and treatment of digestive cancers during COVID-19 in Japan: A Cancer Registry-based Study on the Impact of COVID-19 on Cancer Care in Osaka (CanReCO). PLoS One. 2022;17:e0274918. doi: 10.1371/journal.pone.0274918.

21. Ge L, Gon Y, Kimura Y, et al. Altered mental status in cancer-related emergency hospitalizations: prevalence and recorded reasons for admission. BMC Emerg Med. 2026 Jul 6. doi: 10.1186/s12873-026-01667-9.

22. Yamana H, Moriwaki M, Horiguchi H, Kodan M, Fushimi K, Yasunaga H. Validity of diagnoses, procedures, and laboratory data in Japanese administrative data. J Epidemiol. 2017;27:476–482.

23. Amin MB, Edge S, Greene F, et al. AJCC cancer staging manual. 8th ed. Chicago (IL): Springer; 2017.

24. Brierley JD, Gospodarowicz MK, Wittekind C, editors. TNM classification of malignant tumours. 8th ed. Oxford (UK): Wiley Blackwell; 2017

25. Takashima N, Arima H, Kita Y, et al. Incidence, Management and Short-Term Outcome of Stroke in a General Population of 1.4 Million Japanese-Shiga Stroke Registry. Circ J. 2017;81:1636–1646. doi: 10.1253/circj.CJ-17-0177.

26. White IR, Royston P, Wood AM. Multiple imputation using chained equations: Issues and guidance for practice. Stat Med. 2011;30:377–99. doi: 10.1002/sim.4067.

27. Komlodi-Pasztor E, Gilbert MR, Armstrong TS. Diagnosis and Management of Stroke in Adults with Primary Brain Tumor. Curr Oncol Rep. 2022;24:1251–1259. doi: 10.1007/s11912-022-01280-6.

28. Yamamoto T, Suzuki H, Sakairi Y, et al. Thrombus formation at the stump of the pulmonary vein after lobectomy: a prospective multi-institutional study. Surg Today. 2023;53:1343–1351. doi: 10.1007/s00595-023-02692-y.

29. Douketis JD, Spyropoulos AC, Kaatz S, et al. Perioperative Bridging Anticoagulation in Patients with Atrial Fibrillation. N Engl J Med. 2015;373:823–33. doi: 10.1056/NEJMoa1501035.

30. Innes SC, Yin L, Finnerty GT. Stroke in adults with primary intracranial tumours. J Neurol. 2025;272:212. doi: 10.1007/s00415-025-12929-0.

31. Campello E, Ilich A, Simioni P, Key NS. The relationship between pancreatic cancer and hypercoagulability: a comprehensive review on epidemiological and biological issues. Br J Cancer. 2019;121:359–371. doi: 10.1038/s41416-019-0510-x.

32. Weeks KS, Herbach E, McDonald M, Charlton M, Schweizer ML. Meta-Analysis of VTE Risk: Ovarian Cancer Patients by Stage, Histology, Cytoreduction, and Ascites at Diagnosis. Obstet Gynecol Int. 2020;2020:2374716. doi: 10.1155/2020/2374716

33. Wang JG, Geddings JE, Aleman MM, et al. Tumor-derived tissue factor activates coagulation and enhances thrombosis in a mouse xenograft model of human pancreatic cancer. Blood. 2012;119:5543–52. doi: 10.1182/blood-2012-01-402156.

34. Sasano T, Gonzalez-Delgado R, Muñoz NM, Podoplanin promotes tumor growth, platelet aggregation, and venous thrombosis in murine models of ovarian cancer. J Thromb Haemost. 2022;20:104–114. doi: 10.1111/jth.15544.

35. Hisada Y. Dysregulated hemostasis in acute promyelocytic leukemia. Int J Hematol. 2024;119:526–531. doi: 10.1007/s12185-024-03708-0.

36. Hisada Y, Kawano T, Archibald SJ, Welch JS, Reeves BN, Mackman N. Tissue factor activates the coagulation cascade in mouse models of acute promyelocytic leukemia. Blood Adv. 2023;7:5458–5469. doi: 10.1182/bloodadvances.2023010466.

37. Falanga A, Marchetti M. Cancer-associated thrombosis: enhanced awareness and pathophysiologic complexity. J Thromb Haemost. 2023;21:1397–1408. doi: 10.1016/j.jtha.2023.02.029.

38. Giustozzi M, Proietti G, Becattini C, Roila F, Agnelli G, Mandalà M. ICH in primary or metastatic brain cancer patients with or without anticoagulant treatment: a systematic review and meta-analysis. Blood Adv. 2022;6:4873–4883. doi: 10.1182/bloodadvances.2022008086.

39. Turner M, Murchie P, Derby S, et al. Is stroke incidence increased in survivors of adult cancers? A systematic review and meta-analysis. J Cancer Surviv. 2022;16:1414–1448. doi: 10.1007/s11764-021-01122-7.

40. Terada H, Nakamura K, Fujita S, et al. Incidence and risk factors for ischemic stroke in patients with cancer: A retrospective observational study. Thromb Res. 2025;254:109455. doi: 10.1016/j.thromres.2025.109455.

41. Wada S, Yoshimura S, Miwa K, Iwanaga Y, Koga M, Toyoda K. Current status and future aspects in the Japan Stroke Data Bank. Front Neurol. 2023;14:1090136. doi: 10.3389/fneur.2023.1090136.

42. GBD 2021 Stroke Risk Factor Collaborators. Global, regional, and national burden of stroke and its risk factors, 1990-2021: a systematic analysis for the Global Burden of Disease Study 2021. Lancet Neurol. 2024;23:973–1003. doi: 10.1016/S1474-4422(24)00369-7.

